# Deep brain stimulation effects on cortical activity across frequency bands and contact locations

**DOI:** 10.1101/2025.07.18.25331716

**Authors:** Alena Kutuzova, Cosima Graef, Bradley J Lonergan, Aminata Bocum, Yen F Tai, Shlomi Haar

## Abstract

Deep brain stimulation is an effective treatment for Parkinson’s disease, but clinical programming remains subjective and time-consuming. Neurophysiological biomarkers may offer an objective and scalable approach to guide stimulation settings. To examine whether cortical oscillations—particularly in alpha, beta, and narrowband gamma bands—reflect deep brain stimulation parameter changes and their dependency on the stimulating contact location. Thirteen Parkinson’s patients with subthalamic deep brain stimulation (21 hemispheres) underwent electroencephalography during routine programming sessions. Arm-task segments across multiple stimulation amplitudes were analysed. Alpha (8–12 Hz), beta (13–30 Hz), and narrowband gamma (60–90 Hz) power and burst features were extracted from the ipsilateral motor cortex. Volumes of tissue activated were computed and overlapped with the motor subthalamic nucleus to assess stimulation targeting. Relationships between neurophysiological features, stimulation amplitude, motor subthalamic-nucleus overlap, and active contact location were evaluated.

Cortical alpha burst amplitude and duration significantly decreased with stimulation amplitude—but only when active contacts were located within the motor subthalamic-nucleus. Cortical beta-band features showed no significant modulation across amplitudes or locations. Cortical narrowband gamma power and burst rate increased with stimulation amplitude, especially when stimulation overlapped with the motor subthalamic-nucleus, though effects were less spatially specific than for alpha. Cortical alpha and narrowband gamma oscillations provide sensitive and complementary physiomarkers of deep brain stimulation parameter change. Alpha dynamics reflect spatially precise stimulation within the motor subthalamic-nucleus, while narrowband gamma scales with amplitude. These features may support EEG-guided programming and future adaptive deep brain stimulation strategies.

## Introduction

Deep brain stimulation (DBS) is the most widely performed surgical intervention for Parkinson’s disease, offering significant improvements in motor symptoms and quality of life, as well as a reduction in medication requirements.^1^ The procedure involves the implantation of multi-contact electrodes into deep brain structures—typically the subthalamic nucleus (STN) or globus pallidus internus (GPi)—with continuous delivery of high-frequency electrical stimulation. However, in current clinical practice, DBS settings are manually programmed in lengthy and often suboptimal sessions based on trial-and-error adjustments and clinician observation.^2^ This approach contributes to increased patient burden, limits individualization of therapy, and may lead to stimulation-induced side effects such as dyskinesia, speech and mood disturbances.^3^ These limitations underscore the need for a mechanistic understanding of DBS effects and the development of reliable neurophysiological biomarkers that could guide parameter optimization and enable both automated programming^4^ and adaptive, closed-loop DBS systems.^5,6^

### Neurophysiological biomarkers of DBS

Over recent years, multiple electrophysiological approaches and potential biomarkers for DBS—using invasive and non-invasive recordings—were tested.^7^ The most extensively studied neurophysiological biomarkers in Parkinson’s disease is the power of beta oscillations (13–30 Hz), which are pathologically elevated in the basal ganglia and cortex in association with symptom severity—particularly bradykinesia and rigidity—and are suppressed by both dopaminergic medication and effective DBS.^8,9^ Beta-band activity modulation has been identified throughout motor circuits, including the STN^10–12^ and motor cortex.^7,13,14^ However, not all patients demonstrate elevated beta activity, and the correlation between beta power and motor symptom fluctuations remains inconsistent, even within the STN. ^9^

Recent evidence suggests that transient increases in beta activity, known as beta bursts, may serve as a more sensitive and dynamic biomarker of motor impairment in Parkinson’s disease.^15,16^ These bursts, characterized by increased amplitude and duration^17^, are reduced following dopaminergic therapy and during therapeutic DBS.^18^ Importantly, cortical beta power is coherent with subcortical beta activity recorded from the STN^19^ and may be more responsive to changes in DBS parameters.^14,20^ Cortical beta bursts, in particular, have been shown to decrease in amplitude and duration with clinical DBS, correlating with improved motor function.^18^

In addition to beta-band activity, alpha (8–12 Hz) and gamma (particularly narrowband gamma, NBG; 60–90 Hz) oscillations have also been implicated in the modulation of motor and cognitive functions and are dysregulated in Parkinson’s disease. In the alpha range, sensorimotor cortical alpha power has been reported to decrease during STN-DBS, with reductions associated with clinical improvements.^21–23^ A recent study found not only that alpha activity correlates with symptoms at rest, but also that the alpha decrease during movement strongly correlates with the severity of bradykinesia.^24^ Despite these findings, the functional role of cortical alpha oscillations in response to DBS remains poorly understood, and systematic characterization is lacking.

Gamma oscillations, especially within the narrowband gamma range of 60-90Hz, are increasingly recognized for their potential role in facilitating motor performance. Cortical gamma activity has been shown to increase during DBS-induced entrainment and is associated with clinical motor improvements.^25^ Similarly, gamma power and burst rate in the STN are enhanced by dopaminergic medication and correlate with improved movement velocity.^26^ Most prominent NBG findings are focused on finely tuned gamma (FTG), the spectral peak at the subharmonic of the DBS frequency.^20,25,27^ However, the modulation of cortical narrowband gamma when there is no clear FTG, and the NBG bursts modulations by the DBS remain unexplored, representing a key gap in the current understanding of DBS-related cortical dynamics.

### Neuroanatomical stimulation site

The dorsolateral subregion of the STN, commonly referred to as the motor STN (m-STN), is the preferred target for DBS due to its strong connectivity with motor cortical areas and its elevated beta activity in Parkinson’s disease.^28,29^ Anatomical studies have demonstrated that the spatial extent of this beta-oscillatory zone correlates with DBS efficacy, emphasizing the importance of precise electrode placement to optimize therapeutic outcomes.^30–33^ Despite this, many studies investigating neurophysiological responses to DBS do not account for the precise anatomical stimulation location, limiting interpretability and generalizability.

Recent advances in tractography and computational modelling have underscored the potential role of the hyperdirect pathway—linking the STN directly to the motor cortex—in mediating the effects of DBS.^34,35^ Differential activation of this and other white matter tracts, depending on the specific positioning of the stimulation contact, may influence both subcortical and cortical neurophysiological responses. This has important implications for the use of neurophysiological biomarkers, as variability in the stimulation site could confound their reliability and interpretation.

### Study objective

The present study investigated the sensitivity of cortical neurophysiological biomarkers to DBS parameters, and the reliability and practicality of measuring those using non-invasive EEG in a clinical setting. We further examined the influence of stimulation site localization on the relationship between DBS and cortical EEG responses.

## Materials and methods

### Data collection & patient demographics

Data was collected during routine DBS programming sessions at Charing Cross Hospital in London, UK. Participants were selected based on Parkinson’s diagnosis and the presence of DBS leads targeting STN. Participants received an information sheet and were made aware of the requirements of the study before arrival and provided their written informed consent. The study was approved by the local ethics committee (London - Harrow Research Ethics Committee) following the standards set by the Declaration of Helsinki.

A wireless EEG headset with dry electrodes (DSI-7, Wearable Sensing) was used to record 6-channel EEG (F3, C3, P3, F4, C4 and P4 of the 10-20 international system, with Pz as reference and Fz as ground) at 300Hz. This EEG headset enabled rapid set-up without the need for conductive gel and unrestricted movement. Together, these features enabled EEG recording during real-time clinical sessions. EEG data acquisition was continuous throughout the session, during which the clinician adjusted the DBS parameters (contacts, stimulation amplitude, percentage distribution between contacts, frequency, pulse-width) based on their clinical expertise, objective clinical improvement and patients’ subjective symptomatic response. Parkinson’s symptom severity was assessed at the beginning and end of the clinical session using the Unified Parkinsons Disease Rating Scale (UPDRS) part III.^36^ In addition, individual items from the UPDRS-III – which corresponded to the tasks that were used to assess the efficacy of the interim DBS settings, were also scored.

Patients were excluded from the analysis if there were less than 2 valid stimulation setting changes on the same monopolar contact during their session. Valid settings included only monopolar stimulation and a length of task recording of at least 5 seconds. A total of 21 hemispheres from 13 patients with 15 clinical visits were included (Table 1).

**Table 1.** Demographic characteristics of the full patient cohort (All), and subgroups defined by the spatial relationship between the volume of tissue activated (VTA) and the motor subregion of the subthalamic nucleus (m-STN) First level groupings divide patients with VTAs overlapping the m-STN from those whose VTAs did not overlap the m-STN (OFF-Target). Further subdivision of the m-STN group was into those with active contacts located within the m-STN (m-STN IN) or adjacent to it (m-STN ADJ). Three patients (1 female, 2 male) contributed hemispheres to both the m-STN and OFF-Target groups. One patient had active contacts contributing to both m-STN IN and m-STN ADJ subgroups. Three UPDRS scores were missing from the cohort (2 pre-adjustment, 1 post-adjustment), indicated by Ill. Data are presented as median [interquartile range]. Significance between groups in bold indicated with * for *p*<0.05, ** for *p*<0.01 and *** for *p*<0.001.

|  | All | OFF-Target | m-STN | m-STN IN | m-STN ADJ |
| --- | --- | --- | --- | --- | --- |
| Hemispheres (*n*) | 21(11L/10R) | 11(7L/4R) | 13(6L/7R) | 7(3L/4R) | 8(4L/4R) |
| Patients (*n*) | 13(9M/4F) | 9(7M /2F) | 7(4M/3F) | 4(2M/2F) | 4(3M/1F) |
| Patient age (years) | 63 [55.0 68.0] | 57 [55.0 62.3] | 68 [47.3 70.8] | 70 [63.0 73.0] | 55.5 [42.0 67.0] |
| PD duration (years) | 11 [8.25 14.0] | **14 [11.8 17.3]** | **9 [7.3 9]** | 9 [7.0 10.0] | 8.5 [8.0 10.5] |
| DBS duration (years) | 1 [0.23 5.8] | 5 [0.1 6.0] | 1 [0.7 2.9] | 1[0.8 3.5] | 1.8 [0.1 4.3] |
| L-Dopa daily intake (mg) | 687.5 [468.8 1296.9] | **1314.5 [953.1 1350.0]** | **562.5 [375.0 650.0]** | 562.5 [375.0 593.8] | 537.5 [218.8 943.8] |
| Stimulation Amplitude (mA) | 3.1 [1.2 4.3] | 3.6 [0.6 4.9] | 2.6 [0.7 3.7] | 2.6 [2.4 3.5] | 0.5 [0.3 4.2] |
| Pulse-width (μs) | 57.5 [55.0 60.0] | 60 [52.5 60.0] | 50 [45.0 60.0] | 60 [60.0 141.0] | 40 [25.0 54.2] |
| Frequency (Hz) | 130 [125.6 139.0] | 130 [120.5 136.8] | 130 [105.5 137.5] | 130 [55.6 129.4] | 92 [77.3 139.5] |
| Clinic visits (*n*) | 15 | 7 | 9 | 5 | 4 |
| Total DBS settings (*n*) | 114(51R/63L) | 50(17R/33L) | 60(32R/28L) | 21(9R/12L) | 37(22R/15L) |
| Used DBS settings (*n*) | 105(46R/59L) | 46(17R/29L) | 52(26R/26L) | 21(9R/12L) | 31(17R/14L) |
| DBS parameter deltas (*n*) | 133 | 45 | 68 | 36 | 32 |
| UPDRS pre-adjustment | 33 [21.8 38.0] | 31.5 [21.0 34.0] | 37.5 [25.0 40.5] | 33 [17.5 40.5] | 37.5 [29.5 47.0] |
| UPDRS post-adjustment | 15.5 [11.0 20.0] | 12 [10.3 15.8] | 18.5 [9.5 20.0] | 18.5 [13.0 19.5] | 15.5 [7.0 27.5] |
| UPDRS arm-task specific max | 3.0 [1.5 3.0] | 3.0 [1.5 3.0] | 3.0 [1.6 3.0] | 3.0 [1.6 3.0] | 3.0 [2.6 3.3] |
| UPDRS arm-task specific min | 1.0 [0.4 2.0] | 1.0 [0.3 3.0] | 0.5 [0.5 1.9] | 0.5 [0.1 1.8] | 0.5 [0.4 1.9] |

### Neuroanatomical positioning

Pre-operative 3 Tesla T1-weighted MRI scans and post-operative CT scans were obtained for each patient. To obtain refined neuroanatomical location of the DBS electrodes, MRI and CT scans were co-registered using advanced normalization tools with subcortical refine^37^ and the resulting image was normalized and corrected for brain shift^38^ using LEAD-DBS software^39^. DBS leads were localized with the PaCER^40^ algorithm and refined manually for optimal electrode localization. Contact, voltage and voltage distribution across contacts were used to simulate the volume of tissue activated (VTA) by the stimulation using finite element method.^33^ Volume of overlap between the VTA and the motor-STN (m-STN) was calculated as a percentage relative to the m-STN atlas in the Montreal Neurological Institute (MNI) template space.

All DBS settings (*n*=114) trialled by the clinicians across all patients were split based on active contact locations into those whose VTA had overlap with the m-STN (*n*=60) and those who did not (OFF-Target). Then, we further split the m-STN group into those with active contacts inside the m-STN (m-STN IN, *n*=21) and those with active contacts adjacent to the m-STN (m-STN ADJ, *n*=37) (Table 1; Figure 1).

**Figure 1.**
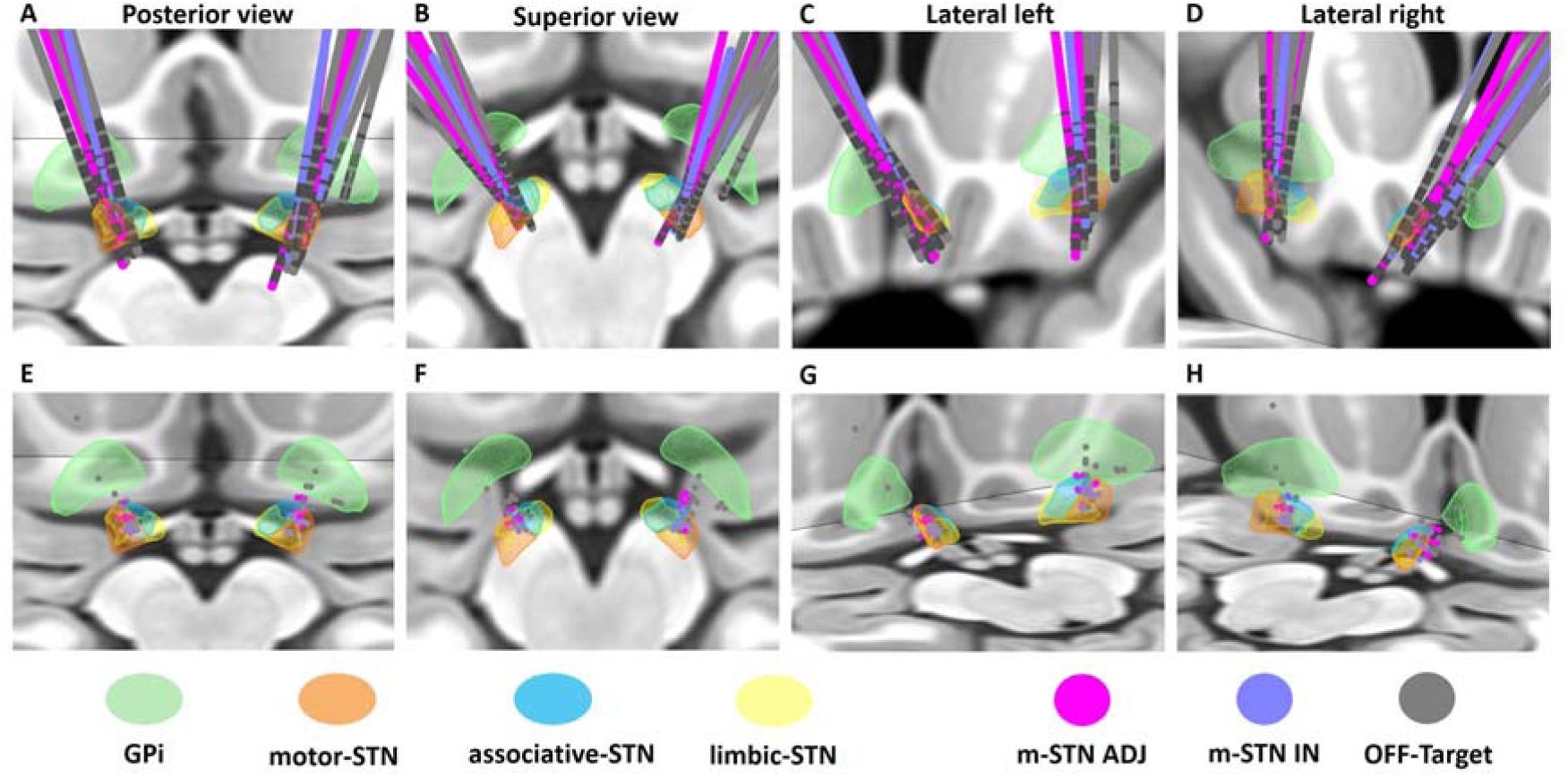
Localization of DBS electrodes and active contacts in MNI space relative to basal ganglia structures. Panels **A–D** show electrode trajectories; panels **E–H** depict active contacts for all patients. Structures of interest include the GPi (green) and subdivisions of the STN: motor (orange), sensorimotor (blue), and limbic (yellow). Views are presented from the posterior **(A, E)**, superior **(B, F)**, left lateral **(C, G)**, and right lateral **(D, H)** perspectives. Panels **A, D, E, G**, and **H** display both axial and coronal slices, whereas panels **B, C**, and **F** show axial slices only. Electrodes are color-coded by target group: blue for those located within the motor STN (m-STN IN), pink for electrodes adjacent to the motor STN (m-STN ADJ), and gray for electrodes outside the STN target (OFF-Target).

### Neurophysiological biomarkers

EEG signals recorded during the clinical session were pre-processed in MATLAB R2023b (The Mathworks, Natick, MA, USA) using EEGLAB^41^. First, the signal was bandpass filtered at 0.5Hz - 98Hz using a 4^th^ order Butterworth filter. Then, the signal was cleaned using EEGLAB’s automated artefact rejection (*clean_rawdata*) and decomposed into an independent component analysis and artefact components were removed with ADJUST, an EEGLAB plug-in for automatic artifact detection.^42^ Finally, EEG segments during arm tasks were extracted for all parameter settings. The selection of arm task segments and not rest segments, as in most studies, was due to the nature of real-world data collection where arm tasks are used in the clinic for assessing the effect of DBS changes on motor outcomes.

For each EEG segment, alpha (8-12Hz), beta (12-30Hz) and NBG (60-90Hz) band oscillatory features were extracted from activity over the motor cortex ipsilateral to the DBS electrode (C3 channel for left DBS and C4 channel for right DBS electrode). Cortical activity over the motor cortex was of interest as it has been shown to have the greatest response to DBS parameters.^14^ To obtain power for each of the bands, we first obtained the power spectral density (*pwelch)* and then identified the most prominent peak per band over the entire clinical session with all DBS settings i.e. all different sets of parameters tested. Then, within the range of the half-prominence of the peak, we identified the power of the most prominent peak for each individual DBS setting. If no peak was found for a particular setting, we used the power at the location of the average peak.

To identify bursts, we used a method based on Tinkhauser *et al*.^43^. First, the signal was bandpass filtered around the location of the average peak (+/-3Hz) for beta and gamma bands. For alpha, we filtered the entire band regardless of the location of the peak because the alpha band is narrow. Next, homomorphic envelopes with carrier frequencies of α = 7.5 Hz, β = 10.5 Hz, and γ = 24.5 Hz were fitted to the signal to estimate the power in the time domain. Bursts were considered as activity greater than 75% of power derived from the entire clinical session. Bursts were required to be longer than approximately 2 oscillations in a given band.^43^ Therefore, we used 100ms for alpha and beta, and 33ms for gamma. Median burst amplitude and duration, as well as burst rate (number of bursts per second) were extracted in each band for each EEG segment.

### Clinical outcomes

To quantify the effectiveness of DBS settings in reducing bradykinesia, each arm-task segment associated with specific bradykinesia-related tasks from the UPDRS-III (finger tapping, hand movements, finger-to-nose manoeuvre, and pronation-supination) was scored by a trained clinician on a scale from 0 to 4, with higher scores indicating more severe symptoms. If a task was repeated multiple times for a setting, the median score was used.

### Statistical analysis

Spearman’s correlations were computed between neurophysiological features and patient demographic information to identify potential covariates. To account for variability across contacts, hemispheres, and patients—and to maximise statistical power—we analysed changes between stimulation settings rather than absolute values. Specifically, we focused on changes in stimulation amplitude and percentage voltage distribution between contacts, while pulse width, frequency, and active contact location were held constant within each pair of settings (delta pairs). These parameters were chosen as they were the most commonly adjusted in our dataset and are known to influence the shape of the simulated VTA, thereby affecting overlap with target regions. We have also identified patients with the most consecutive changes (>4) in stimulation amplitude or percentage voltage distribution and used them as case studies to showcase the efficacy of our group analysis on a patient-specific level.

Robust linear regression models with 95% confidence intervals were used to examine relationships between stimulation amplitude, overlap volume with the m-STN, and neurophysiological biomarkers. Multiple comparisons across spectral features (power and bursting) were corrected using the false discovery rate (FDR). Clinical outcomes were compared across DBS settings using t-tests, restricted to identical task conditions. Statistical significance was set at p < 0.05.

## Results

### Patient Demographics and Clinical Outcomes

Analysis of patient demographics revealed statistically significant differences only between the OFF-Target and m-STN groups, with the OFF-Target group exhibiting higher daily L-Dopa intake and longer disease duration compared to those with m-STN overlap (Table 1). We observed a large difference between pre- and post-adjustment total UPDRS scores in all groups regardless of active contact location. Yet, evaluation of clinical bradykinesia outcomes based on arm-task-specific UPDRS-III scores demonstrated limited sensitivity to subtle changes induced by varying DBS settings. Among all available DBS settings, 79.1% of cases exhibited no change (delta of 0) in arm-task-specific UPDRS-III scores. Stratification based on the degree of m-STN overlap revealed that 76.4% of cases with overlap had no change in scores, compared to 71.4% of OFF-Target cases. Further subgroup analysis showed that within the m-STN overlap group, active contacts located inside the m-STN (m-STN IN) had a zero delta in 66.7% of cases, while those with active contacts adjacent to but still stimulating the m-STN (m-STN ADJ) had a zero delta in 85.7% of cases.

### Alpha-Band Neurophysiological Features

We evaluated the impact of stimulation amplitude on alpha-band neurophysiological features without differentiating by anatomical location and found no significant changes (Figure 2A– D). However, when active contacts were categorized by whether their VTA overlapped with the motor subthalamic nucleus (m-STN), significant differences emerged. Patients with m-STN VTA overlap demonstrated significant reductions in alpha burst amplitude and duration (both *p* < .01, FDR corrected), accompanied by a significant increase in alpha burst rate (*p* < .05, FDR corrected) as stimulation amplitude increased (Figure 2E–H). Further subgroup analysis revealed that these reductions in alpha burst amplitude and duration were driven primarily by contacts located within the m-STN (m-STN IN; both *p* < .05, FDR corrected), whereas no similar relationship was observed in adjacent contacts (m-STN ADJ) (Figure 2J– K). Correlation analyses further reinforced these findings, indicating significant decreases in alpha burst amplitude and duration exclusively in the m-STN IN group as the extent of m-STN overlap increased (both *p* < .05, FDR corrected; Figure 2N–O).

**Figure 2.**
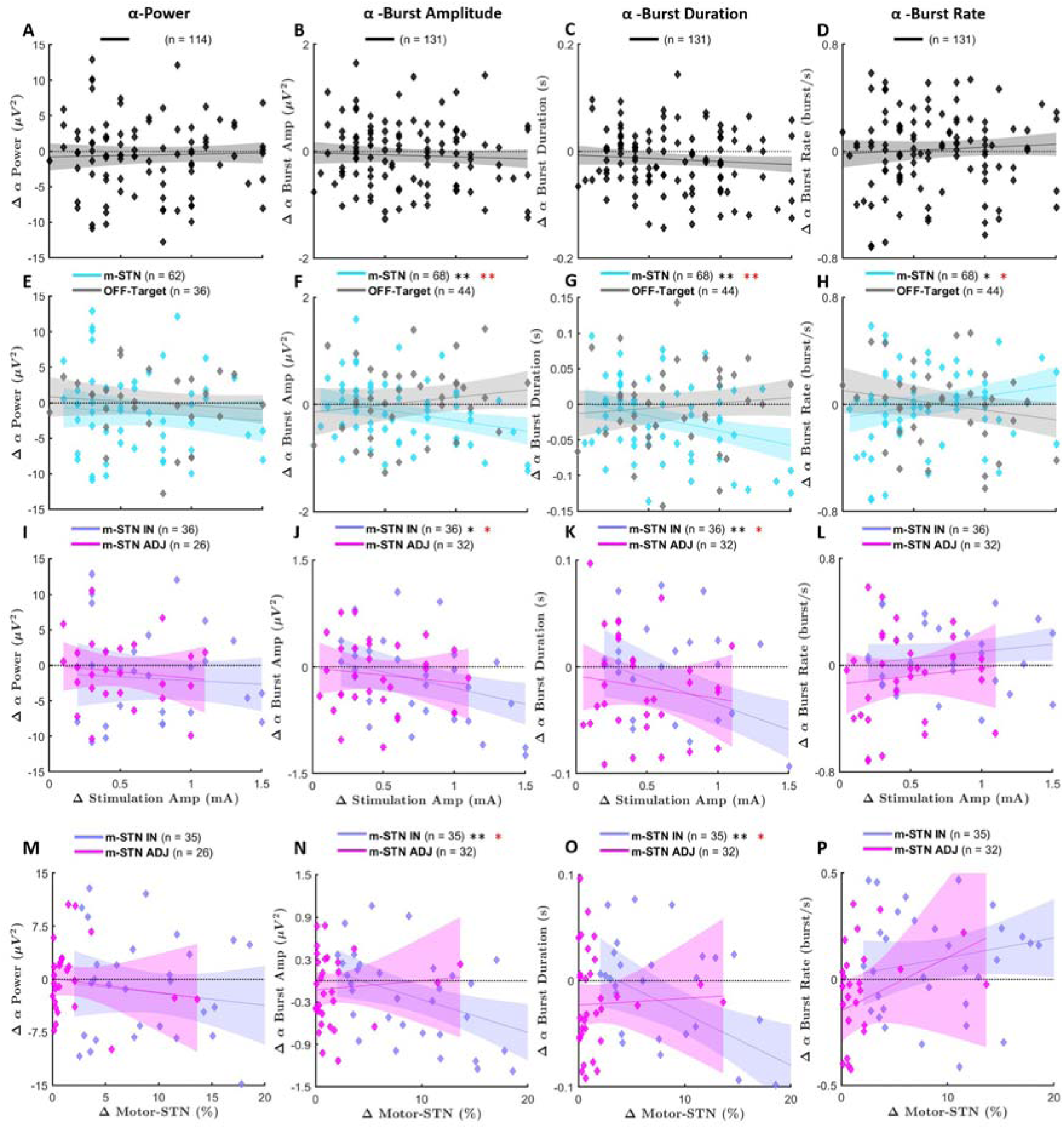
Robust linear regression models depicting the relationship between increases in stimulation amplitude and changes in alpha-band power (A, E, I), burst amplitude (B, F, J), burst duration (C, G, K), and burst rate (D, H, L) Panels **A–D** display data from all participants combined (black). Panels E–H distinguish individuals whose stimulation overlaps with the motor subthalamic nucleus (m-STN, cyan) from those without overlap (OFF-Target, gray). Panels **I–L** further subdivide the overlapping group into patients with active contacts located within the m-STN (m-STN IN, blue) versus those with contacts adjacent to the m-STN (m-STN ADJ, pink). Panels **M–P** illustrate correlations between alpha-band features and the percentage overlap with the m-STN for both m-STN IN and m-STN ADJ groups. Significance indicated with * for *p*<.05, ** for *p*<.01 and *** for *p*<.001 with black for original p-values and red for FDR corrected.

### Beta-Band Neurophysiological Features

No significant relationships were observed between beta-band features and stimulation amplitude across the entire cohort (Figure 3A–D). Additionally, subgroup analyses based on the presence or absence of m-STN overlap did not yield significant correlations between beta-band features and stimulation amplitude (Figure 3E–H). Further stratification into m-STN IN and m-STN ADJ groups similarly showed no significant associations between beta-band features and either stimulation amplitude or the percentage of m-STN overlap (Figure 3I–P).

**Figure 3.**
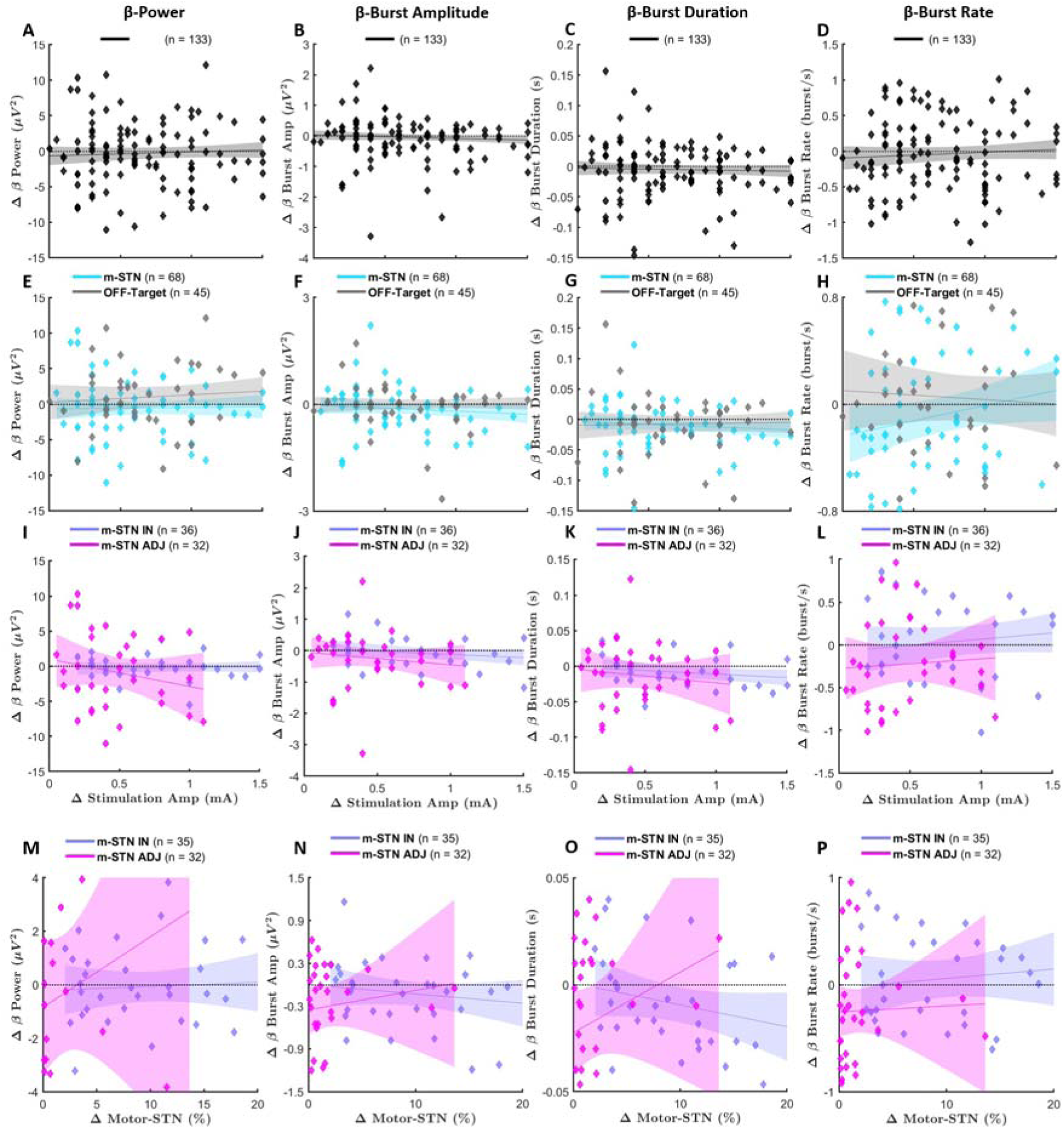
Robust linear regression models depicting the relationship between increases in stimulation amplitude and changes in beta-band power (A, E, I), burst amplitude (B, F, J), burst duration (C, G, K), and burst rate (D, H, L) Panels **A–D** display data from all participants combined (black). Panels **E–H** distinguish individuals whose stimulation overlaps with the motor subthalamic nucleus (m-STN, cyan) from those without overlap (OFF-Target, gray). Panels **I–L** further subdivide the overlapping group into patients with active contacts located within the m-STN (m-STN IN, blue) versus those with contacts adjacent to the m-STN (m-STN ADJ, pink). Panels **M–P** illustrate correlations between beta-band features and the percentage overlap with the m-STN for both m-STN IN and m-STN ADJ groups. Significance indicated with * for *p*<.05, ** for *p*<.01 and *** for *p*<.001 with black for original p-values and red for FDR corrected.

### NBG Features

In contrast to alpha and beta features, NBG features demonstrated significant increases in both NBG power and burst rate with increasing stimulation amplitude across the entire cohort (both *p* < .05, FDR corrected; Figure 4A, D). However, no significant changes were observed in NBG burst amplitude or duration (Figure 4B–C). Dividing the cohort by m-STN overlap revealed that significant increases in NBG power and burst rate were confined to the m-STN overlap group (both *p* < .05, FDR corrected; Figure 4E, H). Further subdivision into active contacts located inside (m-STN IN) versus adjacent (m-STN ADJ) to the m-STN showed that the observed increases in NBG power and burst rate were present only in the m-STN IN subgroup, associated with higher stimulation amplitudes and greater m-STN overlap (Figure 4I, L, M, P). However, these associations did not remain significant after correcting for multiple comparisons.

**Figure 4.**
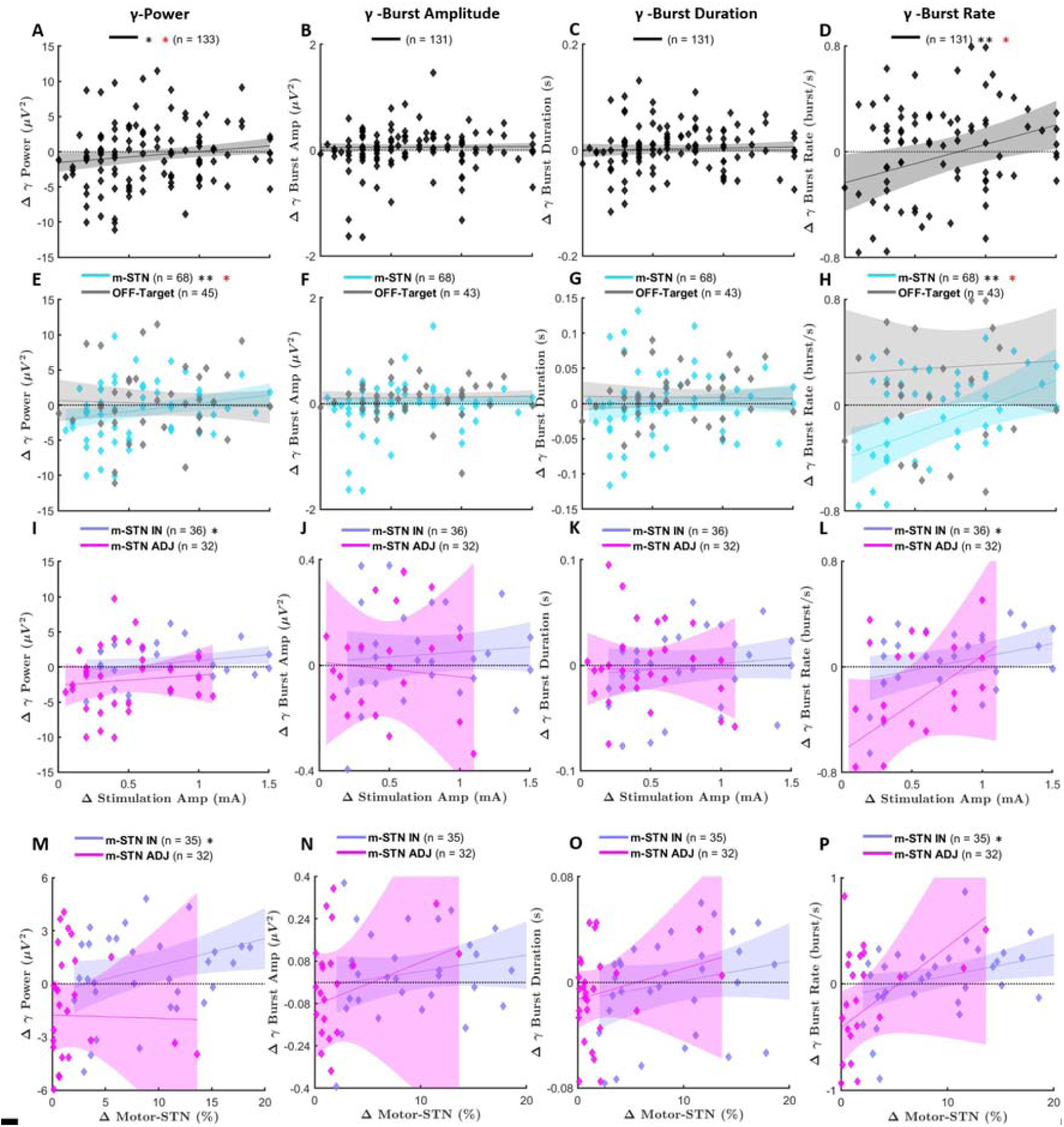
Robust linear regression models depicting the relationship between increases in stimulation amplitude and changes in NBG-band power (A, E, I), burst amplitude (B, F, J), burst duration (C, G, K), and burst rate (D, H, L) Panels **A–D** display data from all participants combined (black). Panels **E–H** distinguish individuals whose stimulation overlaps with the motor subthalamic nucleus (m-STN, cyan) from those without overlap (OFF-Target, gray). Panels **I–L** further subdivide the overlapping group into patients with active contacts located within the m-STN (m-STN IN, blue) versus those with contacts adjacent to the m-STN (m-STN ADJ, pink). Panels **M–P** illustrate correlations between NBG-band features and the percentage overlap with the m-STN for both m-STN IN and m-STN ADJ groups. Significance indicated with * for *p*<.05, ** for *p*<.01 and *** for *p*<.001 with black for original p-values and red for FDR corrected.

### Patient-Specific Analysis

We also investigated the effect of stimulation amplitude on alpha, beta, and NBG features at the patient-specific level (Figure 5). Although statistical significance was not achieved, the patient-specific analysis showed trends consistent with group-level findings (Figure 2E–L). Specifically, when the active contact was within the m-STN (m-STN IN), alpha power, burst amplitude, and burst duration decreased, while the burst rate increased. Conversely, when the active contact was OFF-Target, alpha power decreased, while burst amplitude and duration increased, and burst rate decreased. The m-STN ADJ group exhibited less consistent trends, likely due to the limited availability of consecutive data points.

**Figure 5.**
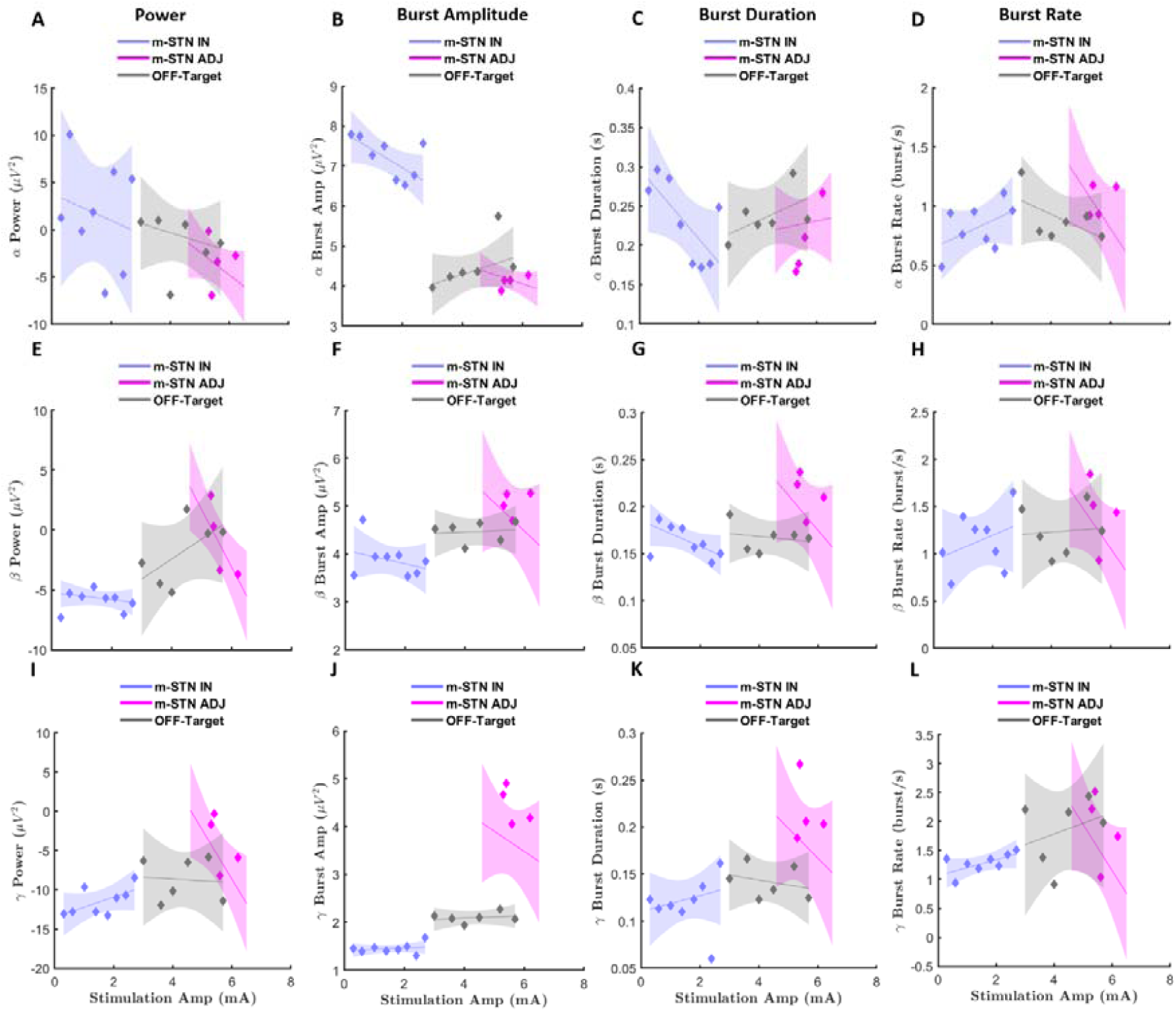
Case studies illustrating the effects of varying stimulation amplitudes on the same active contacts, analysed across alpha (A–D), beta (E–H), and NBG (I–L) features. Panels represent power **(A, E, I)**, burst amplitude **(B, F, J)**, burst duration **(C, G, K)**, and burst rate **(D, H, L)** for three patients, categorized based on the anatomical location of their active contacts: m-STN IN (blue), m-STN ADJ (pink), and OFF-Target (gray). Statistical significance is indicated as follows: \**p* < .05, \*\**p* < .01, \*\*\**p* < .001, with original p-values shown in black and FDR-corrected values in red.

For beta power (Figure 5 E–H), the m-STN IN and OFF-Target groups demonstrated trends similar to the group analysis (Figure 3E–L). Beta power, burst amplitude, and burst duration slightly decreased in the m-STN IN group as stimulation amplitude increased, while burst rate increased. In the OFF-Target group, beta power increased with no significant changes in burst characteristics. In the m-STN ADJ group, a marked decrease in beta power was observed, but changes in burst features remained inconclusive due to limited data.

Lastly, NBG power analysis (Figure 5 I–L) indicated that all NBG features increased with rising stimulation amplitude when the active contacts were within the m-STN (m-STN IN). No consistent trends were observed in the OFF-Target and m-STN ADJ groups. Despite the limited data in some subgroups, these patient-specific analyses support the group-level trends identified earlier in the study.

## Discussion

This study investigated the neurophysiological effects of DBS on cortical oscillations in Parkinson’s disease, focusing on alpha, beta, and NBG oscillatory biomarkers as a function of stimulation amplitude and anatomical targeting within the STN. Using non-invasive EEG recorded during routine clinical programming, we found that: (i) cortical alpha bursts are most sensitive to the accurate placement of the active contact within the m-STN, and attenuate as stimulation amplitude rises and overlap with the m-STN increases; (ii) beta-band metrics are insensitive to small DBS amplitude adjustments and stimulation location; and (iii) NBG power and burst rate scale with stimulation amplitude when the VTA intersects the m-STN, but are less sensitive to the exact active contact location. Together, these results demonstrate that cortical alpha and NBG provide complementary, frequency-specific signatures of stimulation efficacy that are detectable even when clinical UPDRS-III arm-task subscores do not change.

### Alpha oscillations signal anatomically precise m-STN engagement

Alpha-band activity, though less emphasised than beta in the Parkinson’s literature, has been linked to motor readiness and cortical gating.^21,22^ Here, alpha burst amplitude and duration fell as stimulation amplitude and m-STN overlap increased when active contacts lay inside the m-STN. Alpha burst rate was independent of the location of active contacts but increased with the increase in stimulation amplitude when stimulation overlapped with the m-STN. The location-specific response of alpha burst amplitude and duration extends earlier reports that STN-DBS suppresses sensorimotor alpha power^23^ and suggests that alpha bursts may be a practical EEG marker for optimal DBS parameter setting. Mechanistically, selective alpha attenuation may reflect preferential modulation of the hyper-direct cortico-subthalamic pathway that terminates in the dorsolateral STN.^33,35^

### Beta activity shows limited short-term sensitivity

Beta-oscillation power and bursting in the STN are typically elevated in untreated Parkinson’s and correlate with bradykinesia and rigidity.^8,9^ While previous studies reported similar trends in cortical beta^7,13,14^, our study found no significant beta modulation when increasing stimulation amplitude, even when stratifying by contact location. This may reflect a “ceiling” or “floor” effect at clinically optimized settings where beta suppression is maximal.^15^ Moreover, the use of repetitive arm movements during data collection may have induced task-related beta desynchronisation, potentially masking DBS-related changes.^44,45^

### NBG reflects amplitude-dependent cortical engagement

NBG power and burst rate increased linearly with stimulation amplitude across the cohort, with statistically significant changes observed primarily in patients who have m-STN overlap. Invasive recordings suggest that STN gamma bursts correlate with movement velocity and dopaminergic medication^26^, while cortical gamma entrainment accompanies effective DBS.^25^ Our EEG data supports these findings, highlighting that cortical NBG may serve as a sensitive marker of DBS amplitude. The ability of NBG changes to manifest even when active contacts are adjacent to the m-STN suggests that NBG may capture a more spatially diffuse, amplitude-driven cortical response compared to alpha, indicating that cortical alpha might be a more precise marker for effective DBS.

### Translational implications for automatic and adaptive DBS

The dissociation between clinical arm-task subscores—largely unchanged across amplitude steps—and cortical biomarkers highlights the limited granularity of symptom observation during DBS programming. Real-time EEG measures of alpha and NBG could guide rapid, objective adjustment of stimulation parameters, particularly in patients with DBS devices that lack local field-potential sensing. While beta suppression has been used as a control signal in closed-loop systems^46,47^, incorporating alpha and NBG biomarkers could improve specificity and robustness.

Importantly, we demonstrated that the group trends observed in our study persist at a patient-specific level, indicating that our findings may be directly applicable in clinical settings. This patient-specific consistency supports the potential use of these biomarkers during DBS adjustment sessions, enabling clinicians to make more informed and individualized adjustments to stimulation parameters.

### Limitations and future directions

Several limitations warrant consideration. The modest sample size may have limited power for detecting subtle effects, particularly in subgroup analyses. EEG recordings were obtained during movement tasks, restricting the assessment of resting-state beta activity; future studies should include both rest and task conditions to capture state-dependent dynamics. We used low-density EEG with 6 channels, which limits the range of analyses that can be conducted. Higher-density EEG systems could enable the application of source localization algorithms^14^, providing more detailed spatial resolution of cortical activity, but takes longer to set up and thus cannot be used during a standard clinical visit and would not be translatable to clinical practice. Furthermore, as a non-invasive method, EEG does not directly access subcortical signals. Combining cortical EEG with subcortical local field potential recordings will be essential to understand the mechanisms of DBS-related modulation. Additionally, our findings are limited to STN-DBS. Future studies should explore cortical responses to GPi- and ventralis intermedius nucleus (VIM) DBS to assess target-specific biomarkers. Integrating these EEG features into real-time programming tools or adaptive DBS systems is a promising direction for biomarker-guided neuromodulation.

## Conclusion

This study demonstrates that cortical alpha and NBG oscillations serve as sensitive and complementary markers of DBS parameter changes in Parkinson’s disease. Alpha burst features reflect spatial specificity, being selectively modulated when active contacts are within the m-STN, while NBG activity scales with stimulation amplitude regardless of precise contact location, but is indicative of stimulation overlap with m-STN. In contrast, cortical beta-band dynamics show limited responsiveness to clinical adjustments of DBS. Importantly, we demonstrated that the group trends observed persist at a patient-specific level, suggesting that our findings may be applied to patients during clinical DBS adjustment sessions. These results emphasize the potential of EEG-derived biomarkers to enhance conventional programming, particularly in DBS devices lacking subcortical sensing, and pave the way for more precise, efficient, and personalized neuromodulation strategies.

## Data Availability

Data produced in the present study are available upon reasonable request to the authors

## Acknowledgements

The study was supported by the Edmond and Lily Safra Fellowship awarded to S.H. and by the UK Dementia Research Institute Care Research & Technology Centre.

